# Comparison of mental health and burnout between medical and nonmedical students

**DOI:** 10.1101/2025.06.29.25330527

**Authors:** Valerie Carrard, Céline Bourquin, Sylvie Berney, Pierre-Alexandre Bart, Patrick Bodenmann, Alexandre Berney

## Abstract

The important rates of mental health issues and burnout among medical students have been well established. Some suggest that these high rates are due to medical studies being particularly demanding compared to other undergraduate trainings. However, research comparing mental health and burnout of medical and nonmedical students yields inconsistent findings, and are limited by small sample sizes as well as seldom consideration of potential confounding risk factors. This study aimed to complement past research by comparing the mental health and burnout of medical students to those of the nonmedical students in the same university, while accounting for confounding risk factors. A total of 1057 medical and 870 nonmedical students participated in the study, completing validated questionnaires measuring mental health (depressive symptoms, suicidal ideation, anxiety) and burnout (emotional exhaustion, cynicism, academic efficacy), as well as 14 risk factors pertaining to sociodemographic (incl. deprivation), lifestyle, psychological characteristics, life stress, and social relations. After conducting t-tests to compare both groups, the impact of the risk factors were assessed with adjusted regressions. Results revealed that medical students reported significantly less depressive symptoms, suicidal ideation, anxiety symptoms, and cynicism than nonmedical students. They also presented less of the examined risk factors. After adjusting for these factors, medical students still exhibited lower suicidal ideation and cynicism, but all other differences became non-significant. Moreover, when accounting for risk factors, a new significant difference appeared with medical students presenting *more* emotional exhaustion than nonmedical students. These findings suggest that medical studies are not inherently more taxing than other undergraduate disciplines. Both medical and nonmedical students face significant mental health challenges, likely reflecting the typical strains of young adulthood, exacerbated by the pressures of demanding studies. The results underscore the need for holistic interventions to support the mental wellbeing of all undergraduate students, regardless of their discipline or faculty.

## Introduction

Mental health and burnout of medical students have been a preoccupation in the last decades. The fact that medical students present more mental health issues and burnout than the general population is well documented by reviews and meta-analyses [1–3]. A meta-analysis of meta-analyses indeed reports that 32.5% of medical students suffer from depression, 8.9% have suicidal thoughts, gestures, or acts, 32.5% present anxiety, and 35.8% exhibit burnout [4].

It has been suggested that these mental health issues and burnout might be due to the strains specific to medical studies. Indeed, medical studies are reported to be particularly challenging due to important workload, time pressure, stressful exams, competitive environment, relationship with hierarchy, co-workers, and patients during clinical rotations, as well as the confrontation with uncertainty, suffering, and death [5]. To test whether the difficulties of medical studies indeed triggers more mental health issues and burnout, several studies compared prevalences of mental health issues and burnout between medical and nonmedical students. However, the results are mixed. Some studies found that medical students present more depression [6,7] or burnout [6] than nonmedical students (physical education students or other college graduates), but a number of studies found no differences in depression [8–12], suicidal ideation [9,10], or anxiety [3,7] (compared to humanities, nursing, pharmacy, physical education, or business students), or that medical students present actually less depression [13–17] or anxiety [8,17,18] than nonmedical students (humanities, non-medical life-sciences, nursing, health sciences, architecture, English, law, or business students).

To shed light on the question, more studies in a single university setting, comparing more than two disciplines or faculties, and with bigger sample size are needed. Also, it is essential to keep in mind that mental health and burnout are complex phenomenon that can be influenced by numerous risk factors. Most of past studies comparing mental health or burnout of medical students to nonmedical students included gender and sometimes curriculum stage as control variables in order to isolate the specific impact of medical studies [3,6–18]. However, Sheldon’s et al. [19] systematic review and meta-analysis of 66 longitudinal studies identified numerous other factors that significantly impact different indicators of students’ mental health (depression, suicidal ideation, and other psychological distress indicators). Using Furber’s validated taxonomy of mental illnesses’ risk factors [20], they grouped them under 7 categories: *sociodemographic* (e.g., gender, socio-economic status), *physiological and health* (e.g., illness, sleep), *lifestyle* (e.g., physical activity), *psychological* (e.g., personality), *predictors of response to trauma* (e.g., additional life stress), *relational* (e.g., social support), and *factors related to higher education* (e.g., academic environment). Importantly, these risk factors possibly differ between the medical and nonmedical student populations. To ascertain that it is the specificities of the studies that explain the potential differences in terms of mental health and burnout between medical and nonmedical students, research needs to also test the concurrent impact of these non-academic factors that are known to put student at risk for mental health issues and burnout and that might differ between the medical and nonmedical students’ population.

Therefore, the present study proposes to complement the existing literature by testing in a big sample whether medical students differ from all students in other faculties (i.e. social and political sciences, law, humanities, business, geosciences, pharmacy, biology, and theology students) within a single university. The first research question we strive to answer was thus: *do medical students differ from nonmedical students in terms of mental health (depressive symptoms, suicidal ideations, anxiety symptoms) and burnout (emotional exhaustion, cynicism, academic efficacy)?* Also, while it is virtually impossible to account for all the potential mental health risk factors identified in past literature, adjusting the analyses only for gender and curriculum stage as it has been done previously seems too limited to understand the impact of medical studies on such complex and multifaceted concept that is mental health or burnout. Thus, the second research question of the present study was: *are the differences between medical and nonmedical students in terms of mental health and burnout due to the specificities of the studies or to other risk factors?* For the other risk factors, we used the classification proposed by Sheldon and included sociodemographic, physiological, lifestyle, psychological, trauma-related, and relational risk factors.

## Methods

### Design and participants

This cross-sectional observational study is part of the ETMED-L project [21], a larger longitudinal project running from 2020 to 2024 that examines medical students’ interpersonal competences and mental health with an open cohort design.

#### Medical students

As part of the ETMED-L project, an online questionnaire [22] was sent once a year to all medical students at the University of Lausanne (Switzerland), except external exchange students. The questionnaire took approximatively one hour, and the students received 50CHF (≅50USD) for each completed questionnaire. The Lausanne Medical School is a six-year curriculum. Students from all six curriculum years were invited to participate, except in the last data collection in which first-year students were excluded to favor longitudinal participation. Thus, the present study used the ETMED-L data of the 2022 questionnaire for the first-year medical students and that of the 2023 questionnaire for all other curriculum year (2 to 6). Each year the questionnaires were filled in from November 1 to December 2.

#### Nonmedical students

On the 1^st^ of March 2024, a reduced version of the ETMED-L questionnaire was sent by email to all students from all faculties of the University of Lausanne, except the medical faculty (i.e. social and political sciences, law, humanities, business, geosciences, pharmacy, biology, and theology faculties). The students had until April 1 to fill in the questionnaire. Note that the medical faculty is the only one with a six-year curriculum in Lausanne, all the other have a five-year curriculum and all nonmedical students from all five curriculum years were invited to participate. The reduced questionnaire for the nonmedical students took approximatively 15 minutes to complete, and the students received no compensation for their participation.

The Research Ethics Committee of the Canton de Vaud approved the ETMED-L project, including the nonmedical student’s data collection (project number 2020-02474) and all participants gave written informed consent.

### Measures

#### Mental health and burnout

The validated French version of well-established instruments measuring three indicators of mental health and three dimensions of burnout were used in the present study. For mental health, *depressive symptoms* were measured with the Center for Epidemiological Studies-Depression (CES-D) [23,24]. A 20-item instrument assessing the occurrence of symptoms associated with depression over the past week. The CES-D showed a reliable prediction of depression among university students using cut-offs scores of 16 for males, 20 for females, and 19 overall [25]. *Suicidal ideation* was measured with two items of the Beck Depression Inventory (BDI) assessing hopelessness and suicidal thoughts [26,27]. *Anxiety symptoms* were measured with the trait subscale of the State-Trait Anxiety Inventory (STAI), a 20-item instrument which measures the level of anxiety participants “generally feel” [28,29]. For burnout, the Maslach Burnout Inventory Student-Survey (MBI-SS) was used to assess the three dimensions of *emotional exhaustion* (5 items), *cynicism* (4 items), and *academic efficacy* (6 items, reversed dimension) [30,31].

#### Mental health risk factors

We used Sheldon’s et al. [19] categorization to approach exhaustivity of risk factors considered, and included at least one indicator for each category that was not related to the academic environment. As a result, 14 risk factors were included in the present study. For the *Sociodemographic factors*, we included identifying as male (1 = male, 0 = female or non-binary), curriculum year, and the material deprivation index (i.e. financial and basic necessities deprivation) of the Deprivation in Primary Care Questionnaire [DiPCare-Q; 32]. For the *Physiological and health factors*, we used the health deprivation index (i.e. presence of disabilities and chronic conditions impacting daily life) of the DiPCare [32] and the self-reported number of hours of sleep per night. For *Lifestyle*, the number of hours of physical activities per week was assessed. For the *Psychological* factors, we included coping strategies (emotion-focused, problem-focused, and help-seeking strategies measured separately with the coping section of the Euronet questionnaire [33]) and self-reported satisfaction with health (from 1=“very dissatisfied” to 5=“very satisfied”). For the *Predictors of response to trauma*, which includes negative life events, childhood trauma, and additional life stress [19], we measured the number of hours spent in a paid job (on top of studies) per week as additional life stressor. For the *Relational* factors, we used the social deprivation index (i.e. isolation and lack of social activities) of the DipCare [32] and the availability of social support (emotional and practical support measured separately using an adaptation of the Swiss Household Panel items [34]).

### Statistical analyses

After computing means and standard deviations for the overall, medical, and nonmedical samples, multiple imputation was applied (20 imputations) for the variables presenting an overall 5% of missing values or more using Multivariate Normal imputation. Variables with less than 5% missing data were not imputed, as studies indicate that multiple imputation yields minimal information gain at such low missing rates [35].

Then, independent sample t-tests were applied to examine whether and how the medical students differ from the nonmedical students regarding mental health (depressive symptoms, suicidal ideations, and anxiety symptoms), burnout (emotional exhaustion, cynicism, and academic efficacy), and the mental health risk factors included in the present study. Cohen’s *d*s were computed to estimate effect size with *d*s of 0.2, 0.5, and 0.8 being considered respectively as small, medium, and large [36].

To test the impact of the mental health risk factor on the difference between medical and nonmedical students in terms of mental health and burnout, adjusted regressions with robust standard errors were modelled for each mental health indicator (depressive symptoms, suicidal ideations, and anxiety symptoms) and burnout dimension (emotional exhaustion, cynicism, and academic efficacy) separately with the samples (medical vs nonmedical students) as independent variable and the 14 mental health risk factors as covariates. Standardized betas were reported with values of .10-.29 being considered as small, .30-.49 as medium, and .50 or greater as large [36]. According to the commonly recommended guideline of having at least 10 observations per independent variable in a regression model, a minimum sample size of 150 is required to ensure adequate statistical power in the present study.

Additional sensitivity analyses were run. First, the t-tests and adjusted regressions were rerun with the complete non-imputed cases only to identify any potential impact of the multiple imputation applied. Then, given that the Lausanne Medical School is a six-year curriculum, whereas the other faculties have a five-year curriculum, the potential influence of this difference was tested by replicating the analyses while excluding the sixth-year medical students.

P-values of .05 were considered significant and all analyses were run with Stata 18 [37].

## Results

### Sample

Among the 1988 eligible medical students, 1138 participated and 1093 (53.17%) were included in the present study after exclusion of 45 students who gave a wrong answer to the attention questions placed in the questionnaire. For the nonmedical students, 882 of 11536 eligible students participated and 870 (7.54%) were included in the study after exclusion of 12 students who gave a wrong answer to the attention question (316 from social and political sciences, 158 from law, 155 from humanities, 82 from business, 79 from geosciences, 71 from pharmacy or biology, and 9 from theology). Among this total of 1963 included students (1093 medical and 870 nonmedical students), 1777 were complete cases and 186 contained missing data in at least one of the variables of interest (see Table 1). A Little’s test indicated that the missing data could be considered as completely random in their occurrence (Chi2 distance = 98.24, p = .137). After multiple imputation, missing values were reduced to 1.9%: only 5 medical students and 32 nonmedical students had still missing values on some of the non-imputed risk factors and were excluded from the analyses. The final analyzed sample was thus 1926 with 1088 medical students and 838 nonmedical students.

**Table 1.** T-tests comparing medical students to nonmedical students.

|  | Overall sample<br>(N=1963) |  |  | Medical students<br>(N = 1093) |  | Nonmedical students<br>(N = 870) |  | t-tests<br>(N after MI = 1926) |  |  |
| --- | --- | --- | --- | --- | --- | --- | --- | --- | --- | --- |
|  | %Missing | Mean | SD | Mean | SD | Mean | SD | t | p | Cohen's d |
| **Mental Health** |  |  |  |  |  |  |  |  |  |  |
| *Depressive symptoms* | 8.15 | 20.16 | 11.63 | 18.99 | 11.35 | 21.90 | 11.84 | -5.35 | <.001 | 0.25 |
| *Suicidal ideation* | 8.41 | 0.88 | 1.20 | 0.65 | 1.03 | 1.23 | 1.35 | -10.37 | <.001 | 0.50 |
| *Anxiety symptoms* | 9.02 | 47.56 | 11.97 | 45.98 | 11.92 | 49.97 | 11.65 | -7.24 | <.001 | 0.34 |
| **Burnout** |  |  |  |  |  |  |  |  |  |  |
| *Emotional exhaustion* | 9.32 | 16.56 | 5.19 | 16.50 | 4.91 | 16.66 | 5.59 | -0.79 | .460 | 0.03 |
| *Cynicism* | 9.32 | 10.22 | 4.65 | 9.48 | 4.29 | 11.36 | 4.95 | -8.67 | <.001 | 0.41 |
| *Academic efficacy* | 9.32 | 23.94 | 4.67 | 24.00 | 4.53 | 23.85 | 4.90 | 0.78 | .471 | 0.03 |
| **Risk factors** |  |  |  |  |  |  |  |  |  |  |
| *Identifying as male* | 0.00 | 0.28 | 0.45 | 0.32 | 0.47 | 0.23 | 0.42 | 4.53 | <.001 | 0.20 |
| *Age* | 0.10 | 22.56 | 3.76 | 22.09 | 3.03 | 23.14 | 4.46 | -6.40 | <.001 | 0.28 |
| *Material deprivation* | 1.32 | 1.13 | 1.55 | 1.01 | 1.43 | 1.30 | 1.68 | -4.09 | <.001 | 0.19 |
| *Health deprivation* | 1.88 | 0.30 | 0.56 | 0.23 | 0.51 | 0.40 | 0.62 | -6.76 | <.001 | 0.30 |
| *Sleep hours per day* | 0.25 | 7.12 | 0.94 | 7.03 | 0.92 | 7.22 | 0.97 | -4.41 | <.001 | 0.20 |
| *Physical activities* | 0.25 | 3.42 | 2.94 | 3.51 | 2.86 | 3.31 | 3.02 | 1.49 | .138 | 0.07 |
| *Satisfaction with health* | 0.25 | 3.61 | 0.99 | 3.73 | 0.97 | 3.45 | 0.99 | 6.21 | <.001 | 0.29 |
| *Emotion-focused coping* | 9.37 | 10.12 | 4.14 | 9.57 | 3.99 | 10.96 | 4.23 | -6.98 | <.001 | 0.34 |
| *Problem-focused coping* | 9.37 | 7.17 | 1.77 | 7.27 | 1.69 | 7.02 | 1.87 | 2.99 | .003 | 0.14 |
| *Help-seeking coping* | 9.48 | 5.33 | 2.83 | 5.57 | 2.88 | 4.96 | 2.71 | 4.46 | <.001 | 0.22 |
| *Hours in paid job per week* | 0.15 | 4.22 | 7.53 | 2.96 | 6.62 | 5.79 | 8.28 | -8.44 | <.001 | 0.38 |
| *Social deprivation* | 1.88 | 0.50 | 0.76 | 0.45 | 0.70 | 0.57 | 0.81 | -3.54 | <.001 | 0.16 |
| *Emotional social support* | 9.48 | 8.48 | 2.04 | 8.62 | 1.98 | 8.27 | 2.12 | 3.51 | <.001 | 0.17 |
| *Practical social support* | 9.48 | 7.67 | 2.32 | 7.83 | 2.31 | 7.43 | 2.32 | 3.54 | <.001 | 0.17 |

Note. MI = Multiple imputation. Cohen's *ds* of 0.2, 0.5, and 0.8 are considered respectively as small, medium, and large [36].

### Differences between medical and nonmedical students

The t-tests results displayed in Table 1 showed that medical students reported significantly less mental health issues in all three indicators analyzed with small effect sizes for depressive symptoms and anxiety symptoms, and medium effect size for suicidal ideation. According to the CES-D cutoffs, 47.59% of the medical students are at risk of depression, whereas 55.19% of the nonmedical students can be considered at risk. Regarding burnout, the medical students reported also significantly less cynicism with small effect size but did not differ from the nonmedical students in terms of emotional exhaustion nor academic efficacy.

Overall, medical students also presented significantly less mental health risk factors than nonmedical students. First, they significantly differed in their gender identification with the proportion of students identifying as male being higher in the medical students’ sample (32.20%) than in the nonmedical students’ sample (22.99%). Also, the medical students were younger, presented less material deprivation, reported less health deprivation, slept less hours per night, had higher satisfaction with their health, used more adaptive coping strategies (less emotion-focused, more problem-focused, and more help-seeking coping), and reported having less social deprivation as well as more emotional and practical social support available than nonmedical students. All these differences were significant with p values <.01, but Cohen’s *d*s indicated small effect sizes at best.

### Impact of mental health risk factors

The regressions’ results displayed in Table 2 shows that adjusting for risk factors modifies substantially the differences between medical and nonmedical students in terms of mental health and burnout. Regarding mental health, differences in depressive and anxiety symptoms were largely accounted for by the included risk factors. Specifically, the effect size for depressive symptoms became negligible (β = 0.04), and the difference in anxiety symptoms was no longer significant after adjustment. Suicidal ideation was the only mental health indicator that remained significantly associated with the type of studies, with medical students reporting less suicidal ideation. However, the effect size decreased from medium to small after controlling for the risk factors. For burnout, cynicism remained significant with a small effect size, suggesting that medical studies are associated with lower levels of cynicism compared to nonmedical studies, independent of other mental health risk factors. Furthermore, differences in emotional exhaustion that were otherwise not significant became significant when mental health risk factors are adjusted for, with medical students presenting more emotional exhaustion with small effect size. The same is also observed for academic efficacy (reversed dimension of burnout) with medical students presenting significantly less academic efficacy than nonmedical student, but the effect size indicate a negligeable difference.

**Table 2.** Adjusted regressions testing the difference between medical and nonmedical students (N = 1926)

|  | Mental health |  |  |  |  |  |  |  |  | Burnout |  |  |  |  |  |  |  |  |
| --- | --- | --- | --- | --- | --- | --- | --- | --- | --- | --- | --- | --- | --- | --- | --- | --- | --- | --- |
|  | *Depressive symptoms* |  |  | *Suicidal ideation* |  |  | *Anxiety symptoms* |  |  | *Emotional exhaustion* |  |  | *Cynicism* |  |  | *Academic efficacy* |  |  |
| | $\beta$ | SE | p | $\beta$ | SE | p | $\beta$ | SE | p | $\beta$ | SE | p | $\beta$ | SE | p | $\beta$ | SE | p |
| **Medical students** | .04 | 0.42 | .041 | -.12 | 0.05 | <.001 | -.01 | 0.41 | .550 | .10 | 0.22 | <.001 | -.13 | 0.22 | <.001 | -.07 | 0.22 | .001 |
| *Identifying as male* | -.06 | 0.45 | .001 | .03 | 0.06 | .170 | -.07 | 0.45 | <.001 | -.05 | 0.26 | .029 | .06 | 0.25 | .022 | -.03 | 0.25 | .183 |
| *Curriculum year* | -.09 | 0.12 | <.001 | -.04 | 0.01 | .040 | -.07 | 0.12 | <.001 | -.07 | 0.07 | .002 | .14 | 0.07 | <.001 | -.02 | 0.06 | .404 |
| *Material deprivation* | .07 | 0.14 | <.001 | .02 | 0.02 | .398 | .03 | 0.13 | .072 | .06 | 0.08 | .008 | .05 | 0.08 | .084 | .00 | 0.07 | .843 |
| *Health deprivation* | .19 | 0.46 | <.001 | .20 | 0.06 | <.001 | .14 | 0.41 | <.001 | .14 | 0.20 | <.001 | .06 | 0.22 | .019 | -.06 | 0.21 | .023 |
| *Sleep hours per day* | -.10 | 0.21 | <.001 | -.05 | 0.03 | .022 | -.10 | 0.21 | <.001 | -.11 | 0.12 | <.001 | -.01 | 0.11 | .763 | .05 | 0.11 | .015 |
| *Physical activities* | .00 | 0.07 | .986 | .00 | 0.01 | .938 | -.01 | 0.07 | .494 | -.07 | 0.04 | .001 | .01 | 0.04 | .787 | .03 | 0.04 | .165 |
| *Satisfaction with health* | -.18 | 0.22 | <.001 | -.09 | 0.03 | <.001 | -.16 | 0.23 | <.001 | -.14 | 0.12 | <.001 | -.12 | 0.12 | <.001 | .11 | 0.12 | <.001 |
| *Emotion-focused coping* | .39 | 0.06 | <.001 | .24 | 0.01 | <.001 | .49 | 0.05 | <.001 | .31 | 0.03 | <.001 | .25 | 0.03 | <.001 | -.25 | 0.03 | <.001 |
| *Problem-focused coping* | .01 | 0.12 | .701 | -.03 | 0.02 | .244 | -.01 | 0.11 | .399 | -.01 | 0.06 | .744 | -.06 | 0.06 | .006 | .18 | 0.06 | <.001 |
| *Help-seeking coping* | -.08 | 0.08 | <.001 | -.08 | 0.01 | <.001 | -.05 | 0.08 | .014 | -.03 | 0.04 | .163 | -.07 | 0.04 | .005 | .08 | 0.04 | .001 |
| *Hours in paid job per week* | .00 | 0.03 | .982 | .01 | 0.00 | .571 | .00 | 0.03 | .780 | .00 | 0.01 | .983 | .03 | 0.01 | .138 | -.01 | 0.01 | .571 |
| *Social deprivation* | .06 | 0.28 | .002 | .07 | 0.04 | .001 | .02 | 0.28 | .178 | .06 | 0.16 | .013 | -.02 | 0.15 | .331 | .03 | 0.16 | .335 |
| *Emotional social support* | -.06 | 0.13 | .004 | -.06 | 0.02 | .046 | -.04 | 0.12 | .070 | .02 | 0.07 | .581 | -.06 | 0.07 | .055 | .08 | 0.07 | .009 |
| *Practical social support* | -.11 | 0.11 | <.001 | -.11 | 0.02 | <.001 | -.11 | 0.11 | <.001 | -.05 | 0.06 | .078 | -.02 | 0.06 | .541 | .09 | 0.06 | .002 |
| *F* | 144.91 |  |  | 51.27 |  |  | 184.85 |  |  | 57.40 |  |  | 29.32 |  |  | 33.78 |  |  |
| *F's p-value* | <.001 |  |  | <.001 |  |  | <.001 |  |  | <.001 |  |  | <.001 |  |  | <.001 |  |  |
| *R2* | .55 |  |  | .33 |  |  | .57 |  |  | .32 |  |  | .20 |  |  | .22 |  |  |

Notes. Betas of .10-.29 were considered small, .30-.49 medium, and .50 or greater large [36].

### Sensitivity analyses

The t-tests and adjusted regressions replicated with the complete cases showed no differences in any of the results (see S1 and S2 Tables). Results of the analyses excluding the sixth-year medical students showed also no differences to the one including all curriculum years (See S3 and S4 Tables).

## Discussion

Comparing a large sample of medical students to nonmedical students from all other faculties in a single university, the present study confirms that medical students present elevated rates of mental health issues, but further indicates that these high rates are also present among nonmedical students. Indeed, going against the general belief that medical studies are associated with more distress than other disciplines or faculties, our study showed that medical students actually reported significantly less depressive symptoms, suicidal ideations, anxiety symptoms, and cynicism than nonmedical students. Similar results were reported in some past studies showing that medical students present less depression [13–17] and anxiety [8,17,18] than nonmedical students.

However, the results of the present study show that the differences are generally small and that most of them are due to mental health risk factors that do not relate to the studies per se. Indeed, the medical students’ sample presented a higher proportion of individuals identifying as male, less material, health, and social deprivation, more adaptive coping strategies, and better social support. When controlling for these risk factors in order to isolate the specific impact of the studies per se, the differences observed in terms of depressive and anxiety symptoms became negligeable or non-significant. Also, a difference in terms of emotional exhaustion appeared, suggesting that medical studies trigger more emotional exhaustion than nonmedical studies, but this impact can only be seen when adjusting for confounding factors, because it is counterbalanced by lesser risk factors and more protective factors among medical students. In summary, even though the results of the present study indicate some significant differences between medical and nonmedical students in terms of mental health and burnout, it does not indicate that medical studies are more or less taxing than other studies. As indicated by our adjusted regressions’ results, medical students are less at risk for mental health issues and burnout than the other students primarily because they represent a different type of population that is characterized by less biopsychosocial disadvantages.

As already mentioned, past literature is mixed regarding the differences between medical and nonmedical students in terms of mental health and burnout. Some found that medical students present more mental health issues and burnout [6,7], whereas other found the opposite [8,13–18] or no significant difference [3,7–12]. In a sense, the results of the present study align with all this past literature as they indicate that one can find a significant difference in one direction or the other, or even no significant differences, depending on which indicator of mental health and burnout is examined and which risk factors are adjusted for or not. Therefore, future research on mental health among university students need to include several indicators of mental health and account for potential confounding risk and protective factors to develop a comprehensive understanding of the issues at hand.

Many reasons might explain why medical students present systematically less biopsychosocial disadvantages than nonmedical students. Among them, we could cite differences in financial family support that could facilitate studies reputed as more demanding, better educational opportunities among less deprived populations, sociocultural emphasis favoring medicine in less deprived contexts, higher self-esteem among less deprived individuals, less developed social networks among more deprived populations, and admission regulations that could favor less deprived applicants (e.g., requiring citizenship or permanent residency). These different factors seem to have an impact on the overall observed mental health. There is thus a clear need for more services to support students facing biopsychosocial disadvantages and especially for nonmedical students. Most universities, including ours, already propose these kinds of services such as social services for students in precarious situations, psychotherapeutic and spiritual support to care for the students’ psychological health, varied students’ association and social events to increase the social support available to students, as well as on campus sport facilities and healthy food options to favorize healthy living. Regarding the material deprivation issues, university tuitions are already very affordable in Switzerland (i.e. the equivalent of around 1100 USD/year in our university). Reasonable tuitions could be further supplemented by affordable living facilities and scholarships.

Perhaps the most significant implication of this study’s findings is that both medical and non-medical students face considerable mental strain typical of young adults, exacerbated by the additional challenges of demanding undergraduate studies. Indeed, between 48% and 55% of the present study’s students can be considered at risk of depression according to the CES-D validated cutoff. According to a sensitivity/specificity analysis reported elsewhere [38], this would correspond to around 17% of the students presenting an actual clinical depression. These numbers call for a particular attention to the mental health issues of all university students across all disciplines and faculties though easily accessible and affordable psychotherapeutic services.

### Limitations

While the findings of this study provide valuable insights into the mental health and burnout of medical and nonmedical students, several limitations should be considered when interpreting the results. First, the medical students’ data were collected in November, whereas the nonmedical students’ data were collected in March. While both are exam-free months, they still are at different stages of the academic year and in different seasons. However, the medical curricula structure is drastically different from those of other faculties no matter the year-period and we actually found the reverse of the usually observed seasonal effect with the students surveyed in winter showing less mental health issues and burnout than those surveyed in spring. Secondly, the sample of non-medical students demonstrated a significantly lower participation rate and a higher amount of missing data compared to the medical student sample. This discrepancy can be attributed to the ETMED-L project’s affiliation with the Medical School, which primarily targets medical students. The medical students were financially compensated for their participation, while nonmedical students were not, because the survey for nonmedical students was introduced later in the project and was notably shorter than the one completed by medical students. As a result, participation biases can be expected within the nonmedical students’ sample. Indeed, for instance, males are known to be less interested in participating to health research [39] and were thus under-represented in our nonmedical sample. Nevertheless, the size of the nonmedical sample remains large, and the covariates included in the present study enabled to account for the potential participation biases. Third, interpretations of the results related to suicidal ideation should be approached with caution, as it was not the main focus of the present paper and was thus assessed with only two items. These items were not specifically validated as a comprehensive measure of suicidal ideation and may not capture its full complexity. Finally, by accounting for the differences between medical and nonmedical students in terms of sociodemographic, physiological, lifestyle, psychological, life stress, and relational factors, the present study strived to isolate the specific impact of medical studies. However, it is virtually impossible to totally isolate the impact of the studies themselves, because it is impossible to consider every possible other influencing factors. Nevertheless, the present study goes a step further compared to past research that controlled only for gender and sometimes curriculum stage. Giving that it is impossible to isolate the impact of specific studies and also impossible to manipulate it, future research should favor qualitative approaches to explore the specificities of the different studies in order to reach a comprehensive understanding of the impact of specific studies on mental health and burnout.

## Conclusion

The main take away of our study is that mental health and burnout rates are rather high for all university students no matter their disciplines or faculties and that it warrants interventions. Our results indeed indicate that medical studies cannot be considered as more or less taxing than other studies. Services and interventions aiming to improve the mental wellbeing of university students should thus be developed in a holistic way that considers both the complexities of undergraduate training as well as multiple external and internal resources and aggravating factors.

## Data Availability

The data that support the findings of this study are openly available in zenodo at http://doi.org/10.5281/zenodo.15149289

## Structured disclosure

## Acknowledgments

The authors want to thank Professor Giorgio Zanetti, Nathalie Janz, and Fabienne Thévenaz for their help in the ETMED-L project’s data collection.

## Funding/Support

This work is supported by the Swiss National Science Foundation (grant number 10001C_197442). The funders had no role in study design, data collection and analysis, decision to publish, or preparation of the manuscript.

## Competing Interests

The authors have declared that no competing interests exist.

## Other disclosures

The authors used ChatGPT (https://chatgpt.com/) to improve the concision and clarity of the manuscript in some sections. After using this tool, the authors reviewed and edited the content as needed and take full responsibility for the content of the publication.

## Disclaimers

None

## Previous presentations

None

## Supporting information

**S1 Table. T-tests comparing medical students to nonmedical students based on complete cases**

**S2 Table. Adjusted regressions testing the difference between medical and nonmedical students based on complete cases (N = 1,777)**

**S3 Table. T-tests comparing medical students to nonmedical students while excluding sixth-year medical students**

**S4 Table. Adjusted regressions testing the difference between medical and nonmedical students while excluding sixth-year medical students (N=1790)**

## Notes

### Competing Interest Statement

The authors have declared no competing interest.

### Funding Statement

Yes

### Author Declarations

The Research Ethics Committee of the Canton de Vaud approved the ETMED-L project, including the nonmedical student’s data collection (project number 2020-02474) and all participants gave written informed.

